# Rheumatoid Arthritis and Sarcopenia - a Prospective Single Center Cohort Study of Postmenopausal Women

**DOI:** 10.1101/2023.04.20.23288851

**Authors:** Simeon Schietzel, Matthias B. Moor, Flurina Roos, Odile Stalder, Daniel Aeberli

## Abstract

**Objectives:** The individual and socioeconomic burden of sarcopenia in rheumatoid arthritis (RA) is most relevant. However, longitudinal cohort data are scarce.

**Methods:** Prospective, single-center, controlled, observational cohort study of consecutive 124 postmenopausal women, 53 with RA, 71 healthy controls (HC). Low muscle mass and low muscle strengths was defined according to the European working group on sarcopenia in older people 2019 (appendicular lean mass index [ALMI] via dual-energy x-ray absorptiometry < 5.5 Kg/m^2^; handgrip strength via dynamometer < 16 Kg). Linear regression models were calculated including demographic and anthropometric data, comorbidities, and co-medication as confounders.

**Results:** Median age was 63 (IQR 56, 70), follow-up 2.1 (IQR 2.0, 5.3) years. At baseline, median ALMI was 6.2 (IQR 6.0, 6.5) Kg/m^2^ in RA patients, 6.3 (IQR 5.6, 6.9) Kg/m^2^ in HC (p = 0.64) with no difference in rates of low muscle mass (RA 16.2 % vs. HC 15.1 %). In the fully adjusted model, mean change in ALMI per year was -0.05 (95%CI -0.10 to -0.01) Kg/m^2^ in RA patients and 0.00 (95%CI -0.02 to 0.03) Kg/m^2^ in HC resulting in a differential loss of -0.06 (95%CI -0.11 to -0.01) Kg/m^2^ per year (p = 0.027). For RA patients, the adjusted OR of experiencing any loss of muscle mass was 3.98 (95%CI 1.47 to 10.77) compared to HC (p = 0.007). On average, RA patients lost 0.78 % of muscle mass per year. At baseline, low grip strength was seen in 27.3 % of RA patients and in 2.9 % of HC (p = 0.002). In both groups, grip strength did not decline during study period. TNFα inhibitors were associated with less, T-cell inhibition with greater loss of muscle mass. Low mass at baseline, disease duration and disease activity were not associated with loss of muscle mass.

**Conclusion:** Postmenopausal women with RA have a significant risk of accelerated loss of muscle mass over time.

## INTRODUCTION

Sarcopenia has become an increasingly recognized challenge in rheumathoid arthritis (RA). Around 31% of RA patients suffer from sarcopenia, especially in long standing disease.^1^ From non-RA populations it is known that sarcopenia is independently associated with increased mortality,^2^ cardiovascular disease,^3^ falls,^4^ and considerable loss of function and autonomy.^2^ Sarcopenia is defined as a deficit of muscle mass and muscle function.^5^ Gold standards of measurement are dual x-ray absorptiometry (DXA)-derived appendicular lean mass index (in Kg/m^2^ or Kg/BMI) and functional measures like handgrip strength (in Kg or mmHg) or gait speed (m/s). Studies assessing muscle mass in RA however, are often based on bio-impedance, of cross-sectional design or older, not incorporating up to date cut-offs.^1^ Prospective controlled investigations applying gold standard measurements are scarce.

We conducted a prospective controlled study in 53 postmenopausal women with rheumatoid arthritis and 71 healthy controls applying DXA-derived body composition measurements and handgrip strength ascertainments. We determined the rates of low muscle mass, low muscle strength and sarcopenia and investigated both, 2-years trajectories of muscle mass and muscle function as well as associated factors including disease duration, disease activity and different classes of anti-rheumatoid drugs.

## METHODS

### Study population

The present study was conducted as a prospective, single-center, controlled, observational cohort study of consecutive postmenopausal women with rheumatoid arthritis treated at the Department of Rheumatology and Immunology, Bern University Hospital, Bern, Switzerland. For the control group, we recruited healthy postmenopausal female volunteers by locally distributed flyers and advertisement on the internal hospital website.^6^ This study adhered to the Declaration of Helsinki and was approved by the ethics committee of the Canton of Bern (approval #BE165/13). Inclusion criteria were written informed consent, a diagnosis of rheumatoid arthritis that met the American College of Rheumatology criteria, ^7^ age of at least 45 years, a history of menopause (defined as 12 months of absent menstrual cycle) and at least two muscle assessments by DXA. Exclusion criteria for patients and HC were missing muscle parameters, hormone replacement therapy (HRT), anabolic bone therapy, history of fracture, insufficient image quality, invalid measurement, joint prosthesis, lost, inflammatory diseases except for RA, kidney failure, metabolic diseases (diabetes mellitus, hyper- or hypoparathyroidism, hyper- or hypothyroidism), pregnancy and drug abuse. For healthy controls (HC), additional exclusion criteria were a history of bisphosphonate medication and a history of osteoporosis.

Inclusion and exclusion criteria were assessed from data obtained through manual patient chart review. From 876 screened study participants, 557 participants were female. From theses 213 patients were both postmenopausal and at least 45 years of age. Among the remaining participants, 96 were RA patients and 117 were HC. After further exclusions, 53 RA patients and 71 HC were included into the analysis (**Figure 1**).

**Figure 1.**
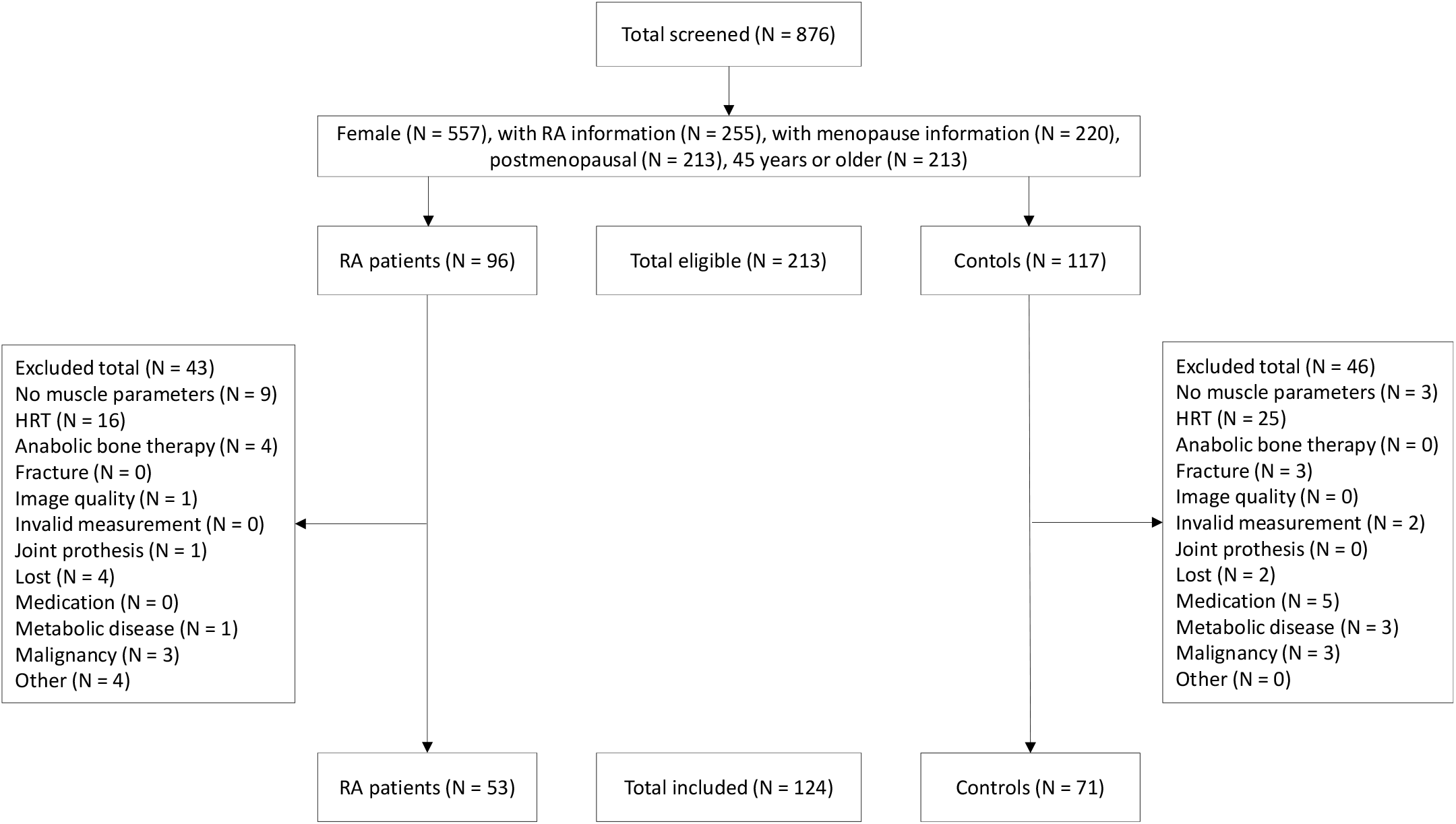
Flow chart screening to inclusion

### Data collection and measurements

#### Clinical parameters

Demographics, comorbidities, medication and blood laboratory data were retrieved from electronic health records. Medication used for the analyses were corticosteroids, vitamin D, biological (biol) DMARD including tumor necrosis factor alpha (TNFα) inhibitors: infliximab, etanercept, golimumab, adalimumab, T-Cell Inhibitors: abatacept, Interleukin-6 receptor antagonist (RA): tocilizumab, B-cell depleting agents: rituximab. Conventional synthetic (cs) DMARDs were methotrexate, hydroxychloroquine, leflunomide and sulfasalazine.

#### Muscle mass

Lean soft tissue mass was measured by whole-body DXA (GE Medical System Lunar Prodigy; GE Medical Systems, Glattbrugg, Switzerland) as reported.^6^ In brief, scans were obtained in array mode at standard supine positioning. Whole-body lean mass was divided in arms, legs and trunk. Body composition was analyzed using enCORE software (version 2004 8.80 001). Appendicular lean mass index (ALMI) was determined as lean mass of all 4 extremities per m^2^. Quality control measures were previously reported.^6^

#### Muscle strength

Handgrip strength was measured as previously described and reported in mmHg. ^6^ In brief, we used a dynamometer (Leonardo GF 8N600591A-S; Novotec Medical GmbH, Pforzheim, Germany) or a soft-tissue dynamometer to measure handgrip strength of the non-dominant hand in averaged triplicates.

#### Definition of low muscle mass, low muscle function and sarcopenia

Low muscle mass, low muscle function and sarcopenia were defined according to the European working group on sarcopenia in older people 2 (EWGSOP2) published in 2019.^5^ Hereby, for women, low muscle mass is defined as an appendicular lean mass index (ALMI) of below 5.5 Kg/m^2^, low muscle strength is defined as grip strengths below 16 Kg. We transformed our values of handgrip strength from mmHg to Kg applying the formula: Handgrip strength (Kg) = 2.991 + 0.113 * handgrip strength (mmHg), according to Martins et al. ^8^ A cut-off below 16 Kg corresponds to mmHg values below 115.1.

### Outcomes

Primary outcome was annual changes in ALMI (Kg/m^2^) and handgrip strength (mmHg) over time in postmenopausal women with RA compared to HC. Pre-specified secondary outcomes included the rate of low muscle mass, low muscle strength and sarcopenia in our population and an analysis of differential decline according to status at baseline. In addition, we formed pre-specified subgroups analyzing rates of muscular decline with regard to DMARDs, disease duration and disease activity (DAS28[ESR]).

### Statistical analyses

All statistical analyses were conducted using Stata version 17.0. Continuous variables are presented with median and interquartile range (IQR) or mean and standard deviation (SD). Categorical variables are presented with number (n) and percentage (%) of patients. All missing data are shown. Characteristics of RA patients and HC were compared using the Chi-squared test or the non-parametric Wilcoxon rank-sum test as appropriate. Baseline was defined as the first visit with available measurements of interest, either ALMI or handgrip.

Outcomes were analyzed through linear mixed-effects models considering all available data, no imputation of the missing values were done. These models include time from the baseline of the patient, RA status and their interaction, the age of the patient at baseline, the time between menopause and baseline. These models were additionally adjusted by different sets of covariates. The list of covariates included age, menopause years, BMI at baseline and vitamin D therapy as time varying variable. Additional analyses of the outcomes for RA patients were only done with linear mixed-effects models. These models include time from baseline, therapy usage and their interaction, the age of the patient at baseline, and the time between menopause and baseline. These models were additionally adjusted by different sets of covariates as indicated.

### Patient and Public involvement

We did not involve patients or the public in the development of the research question or in the design of the study. A link to the published version of the present study will be made available for patients and the public via the website of the Department of Rheumatology and Immunology, Inselspital, Bern University Hospital and University of Bern, Switzerland.

## RESULTS

### Population and baseline characteristics

We included 124 postmenopausal women, 53 with RA, 71 HC. Median follow-up time was 2.1 years (IQR 2.0, 5.3). Median age at baseline was 63 (IQR 56, 70) years and median years since menopause were 13 (IQR 5, 20), both equally distributed between groups (**Table 1**). Rates of vitamin D intake differed significantly between groups (RA 70 % vs. HC 31 %, p < 0.001). For RA patients, disease duration until baseline was 15 (IQR 6, 26) years and disease activity was 2.5 (IQR 2.1, 3.7) according to the DAS28(ESR). 64 % and 62 % of RA patients were on a biological or conventional synthetic DMARD respectively. 26% of RA Patients were upon anti-TNF Therapy as such as infliximab, etancercept, adalimumab or golimumab. Prednisone dose was low with an IQR of 0.0 to 3.5 mg per day (**Table 1**).

**Table 1.** Characteristics of study population at baseline.

|  |  |  | RA (n = 53) | HC (n = 71) | P- value |
| --- | --- | --- | --- | --- | --- |
| **All participants** | Missing - n (%)<br>RA HC |  |  |  |  |
| Age at baseline (yrs) - median (IQR) | 0 (0) | 0 (0) | 65 (58, 70) | 61 (55, 69) | 0.19 |
| Height (cm) - median (IQR) | 2 (4) | 0 (0) | 160 (156, 168) | 166 (162, 169) | 0.003 |
| Weight (kg) - median (IQR) | 2 (4) | 0 (0) | 70 (59, 77) | 63 (58, 72) | 0.11 |
| BMI (kg/m<sup>2</sup>) - median (IQR) | 2 (4) | 0 (0) | 27 (23, 30) | 23 (21, 26) | 0.004 |
| Years since menopause - median (IQR) | 0 (0) | 0 (0) | 14 (8, 21) | 12 (5, 20) | 0.23 |
| Vitamin D therapy - n (%) | 1 (4) | 2 (3) | 37 (70) | 22 (31) | <0.001 |
| **RA patients** | Missing - n (%)<br>RA |  |  |  |  |
| Duration of RA (yrs) - median (IQR) | 1 (4) |  | 15 (6, 26) | N/A |  |
| DAS28 (ESR) (mm) - median (IQR) | 10 (19) |  | 2.5 (2.1, 3.7) | N/A |  |
| Rheumatoid factor - n (%) | 3 (6) |  | 36 (68) | N/A |  |
| Anti-CCP antibody - n (%) | 7 (13) |  | 36 (68) | N/A |  |
| Prednisone (mg/day) - median (IQR) | 0 (0) |  | 0.0 (0.0, 3.5) | N/A |  |
| csDMARD - n (%) | 0 (0) |  | 33 (62) | N/A |  |
| biol. DMARD - n (%) | 0 (0) |  | 34 (64) | N/A |  |
| TNFα inhibitors n (%) | 0 (0) | | 14 (26) | N/A | |
| IL-6 RA - n (%) | 0 (0) |  | 4 (8) | N/A |  |
| B cell depletion - n (%) | 0 (0) |  | 10 (19) | N/A |  |
| T cell inhibition - n (%) | 0 (0) |  | 10 (19) | N/A |  |

Data are presented as number (%) and median (IQR). Only height was normally distributed. For RA patients, normality were found in age (mean 64, SD 9.3 years), height (mean 162, SD 7.4 cm), BMI (mean 27, SD 5.5 Kg/m<sup>2</sup>), DAS28(ESR) (mean 2.9, SD 1.1) and handgrip strength (mean 148, SD 61 mmHg). RA, rheumatoid arthritis; HC, healthy controls; BMI, body mass index; HRT, hormone replacement therapy; DAS28, disease activity score 28; CCP, cyclic citrullinated peptide; csDMARD, conventional synthetic disease-modifying anti-rheumatic drugs; biol. DMARD, biological disease-modifying anti-rheumatic drugs; TNFα , tumor necrosis factor alpha; IL6-RA, Interleukin-6 receptor antagonists.

### Muscle mass and muscle strength

Median (IQR) ALMI at baseline did not differ between RA with 6.2 (6.0, 6.5) Kg/m^2^ and HC with 6.3 (5.6, 6.9) Kg/m^2^ (p = 0.64). RA patients had lower median (IQR) handgrip strength measuring 145 (90, 200) versus HC 220 (180, 270) mmHg (p <0.001) (**Table 2**).

**Table 2.** Muscle parameter at baseline.

|  | Missing |  | RA (n = 62) | HC (n = 73) | P-value |
| --- | --- | --- | --- | --- | --- |
|  | N (%) |  | Median (IQR) |  |  |
|  | RA | HC |  |  |  |
| **ALMI - Kg/m<sup>2</sup>** | 25 (40) | 21 (29) | 6.2 (6.0, 6.5) | 6.3 (5.6, 6.9) | 0.64 |
| **Handgrip strength - mmHg** | 40 (65) | 3 (4) | 145 (90, 200) | 220 (180, 270) | <0.001 |
|  |  |  | N (%) |  |  |
| **Low muscle mass - n (%)**<br>(ALMI <5.5 Kg/m<sup>2</sup>) | 25 (40) | 21 (29) | 6/37 (16) | 8/53 (15) | 1.0 |
| **Low muscle strength - n (%)**<br>(Handgrip <115.1 mmHg ≡ <16 Kg) | 40 (65) | 3(4) | 6/22 (27) | 2/70 (3) | 0.002 |
| **Sarcopenia - n (%)**<br>(low strengths AND low mass) | 56 (90) | 21 (29) | 1/6 (17) | 0/52 (0) | 0.1 |

Sixteen percent of RA patients versus 15 % of HC showed low muscle mass (ALMI < 5.5 Kg/m^2^) at baseline (p = 1.0). Rates of low muscle strength (grip strength < 115.1 mmHg ≌ < 16 Kg) were 27 % in RA versus 3 % in HC (p = 0.002). Rates of sarcopenia (ALMI < 5.5 Kg/m^2^ plus grip strength < 115.1 mmHg) was 17 % in RA patients and 0.0 % in HC (p = 0.1) (**Table 2**).

### Primary outcome – change of muscle mass over time

In the fully adjusted model (n = 120), accounting for age, menopause years, BMI at baseline and vitamin D, mean change in ALMI per year was -0.05 (95%CI -0.10 to -0.01) Kg/m^2^ in RA patients versus 0.00 (95%CI -0.02 to 0.03) Kg/m^2^ in HC. Resulting differential mean loss of muscle mass in RA patients versus HC was -0.06 (95%CI -0.11 to -0.01) Kg/m^2^ per year (p = 0.027) (**Table 3**). In the fully adjusted model (excluding vitamin D due to limited number of events), RA patients had an odds ratio (OR) of 3.98 (95%CI 1.47 to 10.77) for experiencing any loss of muscle mass during the study period compared to HC (p = 0.007).

**Table 3.** Primary outcome. Mean change of muscle mass and muscle strengths per year

| Outcome ** |  | RA | HC |  |  |
| --- | --- | --- | --- | --- | --- |
|  | no.<br>pat.* | Mean change per year<br>(95%-CI) ** |  | Difference in change<br>per year (95%-CI)** | p-value |
| ALMI (Kg/m<sup>2</sup>) | 120 | -0.05 (-0.09 to -0.01) | 0.01 (-0.01 to 0.04) | -0.06 (-0.11 to -0.01) | 0.02 |
| Grip strength (mmHg) | 117 | 2.38 (0.17 to 4.58) | 0.71 (-0.74to 2.15) | 1.67 (-0.90 to 4.24) | 0.203 |

\*nb analyzed patients.
\*\*adjusted for age, menopause years at baseline, BMI and vitamin D.
ALMI, appendicular lean mass index

**Figure 2**. shows the fully adjusted ALMI decrease of RA patients compared to the ALMI of HC, which did not change over the course of the study. The difference in change of ALMI per year between groups was significant (p = 0.027). In RA patients, the average decline per year represents a decrease of 0.86% per year (**Figure 2**).

**Figure 2.**
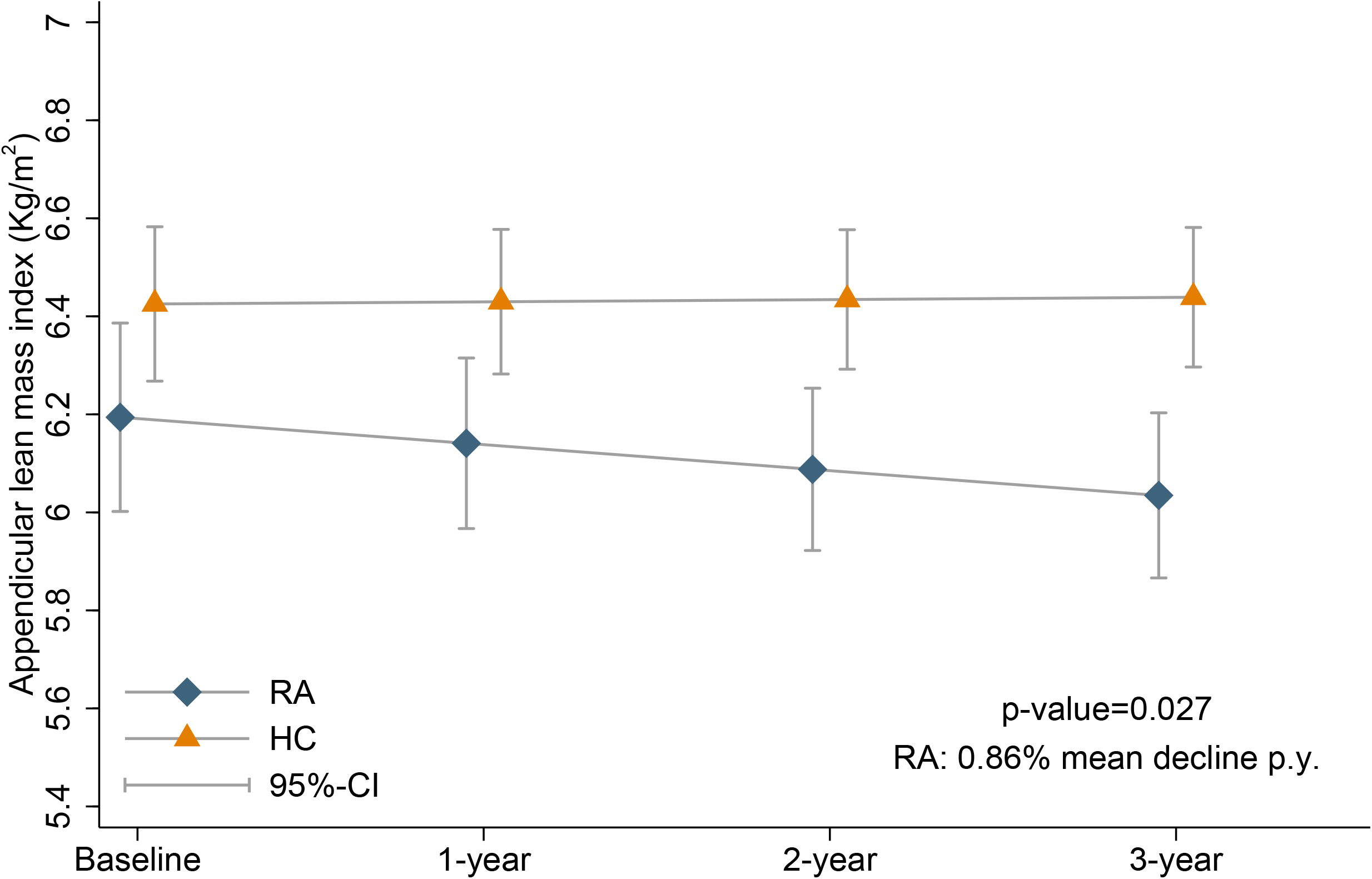
Fully adjusted appendicular lean mass index over time

In the fully adjusted model, RA patients and HC did not lose grip strength over time. RA patients gained 2.38 (95%CI 0.17 to 4.58) mmHg of grip strength per year however, slope difference to HC was not significant (**Table 3**).

### Subgroups Analysis

In the fully adjusted model, loss of muscle mass in RA patients was solely seen in those with normal muscle mass at baseline who lost -0.08 (95%CI -0.15 to -0.01) Kg/m^2^ per year. RA patients with low muscle mass at baseline did not experience a decline in muscle mass showing an annual ALMI change of 0.02 (95%CI -0.04 to 0.08). Difference in annual changes between these groups was significant (0.10 [95%CI 0.01 to 0.19], p = 0.035).

### Annual change of muscle parameters according to DMARDs, disease duration and disease activity

In the fully adjusted subgroup analysis, the use of TNFα inhibitors was significantly associated with sparing of muscle mass over time. Whereas those without TNFα inhibitors lost muscle mass over time (ALMI -0.07 (95%CI -0.13 to -0.02), ALMI did not change during study period in those taking TNFα inhibitors: slope ALMI difference of 0.08 (95%CI 0.02 to 0.15) Kg/m^2^per year in favor of TNFα inhibitors (p = 0.02) (**Table 4**). Grip strength values of patients on csDMARD (methotrexate, hydroxychloroquine, leflunomide, sulfasalazine) did not change compared to those without csDMRD who gained grip strength over time. Difference was -5.59 (95%CI -9.33 to -1.86) mmHg per year (p = 0.003). All other DMARDs were not independently associated with a change in muscle mass or muscle function over time (**Table 4**).

**Table 4.** Secondary outcome. Association of change of muscle mass and grip strength with anti-rheumatic drugs in RA patients

| Outcome ** | no. pat.* | Mean change per year (95%-CI)** |  | Difference in change per year (95%-CI)** | p-value |
| --- | --- | --- | --- | --- | --- |
|  |  | **Prednisone** | **No Prednisone** |  |  |
| ALMI (Kg/m<sup>2</sup>) | 50 | -0.05 (-0.11 to 0.0) | -0.04 (-0.11 to 0.03) | - 0.02 (-0.12 to 0.08) | 0.7 |
| Grip strength (mmHg) | 47 | 2.25 (-.13 to 5.18) | 3.42 (-1.49 to 8.34) | -0.90 (-6.48 to 4.68) | 0.75 |
|  |  | **TNFα inhibitors** | **No TNFα inhibitors** |  |  |
| ALMI (Kg/m<sup>2</sup>) | 50 | 0.01 (-0.03 to 0.05) | -0.07 (-0.13 to -0.02) | 0.08 (0.02 to 0.15) | **0.02** |
| Grip strength (mmHg) | 47 | 0.45 (-4.18 to 5.09) | 3.89 (1.52 to 6.26) | -3.44 (-8.67 to 1.79) | 0.2 |
|  |  | **csDMARD** | **No csDMARD** |  |  |
| ALMI (Kg/m<sup>2</sup>) | 50 | -0.04 (-0.07 to -0.00) | -0.07 (-0.16 to 0.03) | 0.03 (-0.07 to 0.13) | 0.59 |
| Grip strength (mmHg) | 47 | 0.03 (-0.07 to 0.13) | 5.60 (3.19 to 8.01) | -5.59 (-9.33 to -1.86) | **0.003** |
|  |  | **Biol. DMARD** | **No biol. DMARD** |  |  |
| ALMI (Kg/m<sup>2</sup>) | 50 | -0.05 (-0.11 to 0.01) | -0.05 (-0.10 to -0.01) | -0.00 (-0.08 to 0.07) | 0.98 |
| Grip strength (mmHg) | 47 | 2.28 (-0.11 to 4.67) | 5.61 (0.79 to 10.43) | -3.32 (-8.83 to 2.19) | 0.24 |
|  |  | **IL-6 RA** | **No IL-6 RA** |  |  |
| ALMI (Kg/m<sup>2</sup>) | 50 | -0.01 (-0.05 to 0.03) | -0.05 (-0.10 to -0.01) | 0.05 (-0.01 to 0.11) | 0.14 |
| Grip strength (mmHg) | 47 | 2.21 (1.40 to 3.02) | 3.14 (0.47 to 5.80) | -0.92 (-3.70 to 1.85) | 0.51 |
|  |  | **B cell depletion** | **No B cell depletion** |  |  |
| ALMI (Kg/m<sup>2</sup>) | 50 | -0.08 (-0.29 to 0.14) | -0.04 (-0.07 to -0.01) | -0.04 (-0.25 to 0.18) | 0.75 |
| Grip strength (mmHg) | 47 | 3.14 (-0.69 to 6.97) | 2.85 (0.03 to 5.67) | 0.29 (-4.39 to 4.97) | 0.90 |
|  |  | **T cell inhibition** | **No T cell inhibition** |  |  |
| ALMI (Kg/m<sup>2</sup>) | 50 | -0.13 (-0.19 to -0.07) | -0.03 (-0.08 to 0.01) | -0.09 (-0.17 to -0.02) | **0.02** |
| Grip strength (mmHg) | 47 | 0.35 (-1.79 to 2.50) | 3.32 (0.76 to 5.88) | -2.96 (-6.27 to 0.34) | 0.08 |

\* nb analyzed patients.
\*\* adjusted for age, menopause years, BMI at baseline, and vitamin D. For Handgrip strength csDMARD were associated with a significant decline data not shown), all other drugs were not associated with a decline over time. ALMI, appendicular lean mass index, TNFα, tumor necrosis factor alpha; csDMARD, conventional synthetic disease-modifying anti-rheumatic drugs; biol. DMARD, biological disease-modifying anti-rheumatic drugs; IL-6 RA, Interleukin-6 receptor antagonists.

Finally and of interest, neither disease duration, nor disease activity were independently associated with loss of muscle mass or muscle strength over time.

## DISCUSSION

In this prospective, single-center, controlled, observational cohort study of consecutive postmenopausal women we investigated change and predictors of muscle mass and muscle strength over time, in 53 postmenopausal women with RA in comparison to 71 HC.

We found 16.2 % of RA patient to have low muscle mass. Adequate comparisons to the literature is challenging as to the strong association of age and sex, the different methods of measurement, the divergent cut-offs and the numerous correlating factors. We refer to the literature that provides the gold standard of DXA-derived indices of ALMI (in Kg/m^2^ or Kg/BMI) and internationally accepted cut-offs.

Applying almost the same ALMI cut-off (≤ 5.5 Kg/m^2^), a cross-sectional Moroccan study including 107 female RA patients found a considerably higher rate of low muscle mass (37.4 %).^9^ Mean age was considerably younger (52.3 [SD 13.2] vs. 64 [SD 9.3] years in our study), however, RA was more severe (mean [SD] DAS28[ESR] 4.05 [SD 1.39] vs. 2.9 [SD 1.1]) and steroid dose higher (mean 11 [SD 9] vs. 1.9 [SD 2.7] mg/d in our study). A Russian study of 40 postmenopausal RA patients of comparable mean age (63 [SD 7] vs. 64 [SD 9.3] years in our study) also found higher low mass rates (25 %) however, based on the sex-specific lowest 20% of the Health-ABC cohort, ALMI cut-off of was considerably higher (< 5.67 Kg/m^2^).^10^ A Japanese study of 148 female RA patients, applying the Asian cut-off of ALMI (< 5.4 kg/m^2^), also found a higher rate of low mass (25.7 %).^11^ Mean age and disease duration were comparable however, disease activity was higher (mean DAS28[ESR] in normal and low ALMI 3.2 and 4 [SD 1.8] vs. 2.9 [SD 1.1] in our study). A Turkish study (ALMI cut-off of < 5.45 Kg/m^2^) found only slightly higher rates of low mass (20%) in 40 female RA patients despite younger mean age (48.3 [SD 8.3] vs. 64 [SD 9.3] years), higher mean BMI (31.8 [SD 7] vs. 27 [SD 5.5] Kg/m^2^) and shorter disease duration (mean 6.3 [SD 5.7] vs. median 15 [IQR 6, 26] years in our study). Data on disease severity were not given.^12^

Two studies found lower rates of low mass. A Brazilian study of 78 female RA patients, applying a low mass cut-off of ALMI < 5.45 Kg/m^2^, found low mass in only 9%.^13^ From the mixed cohort (men and women, 89 % women), disease severity was comparable (median DAS28[CRP] 3 [IQR 1, 3] vs. 2.5 [2.1, 3.7] in our study), however, mean age was younger (56.5 [SD 7.3] vs. 64 [SD 9.3] years in our study) and median disease duration shorter (8.5 [3, 18] vs. 15 [6, 26] years in our study). A study from New Zeeland applied the low mass definition of the foundation of the national institutes of health (FNIH) in Kg/BMI with a cut-off of < 0.512 Kg/BMI and found comparable low mass rates (15 %) to our study. Age range (61.1 [SD 13.3] vs. 64 [9.3] years in our study) and disease severity (DAS28[ESR] 3.6 [SD 1.3 vs. 2.9 (1.1)] in our study) were comparable.^14^

Summarizing, the rate of low muscle mass in our population (16.2 %) seems to be in the low spectrum compared to the DXA-derived literature on muscle mass in RA (9 - 37.4 %).^9-14^ Possible reasons are low disease severity, low dose and rates of glucocorticoid use as well as the stringent treat to target therapy, favoring glucocorticoid sparing DMARDs and long term remission, as well as the exclusion of multi-morbidity in this population. However, valid comparative analyses are a challenge, especially when crucial confounders like exact age group, years since menopause, nutrition, lifestyle and physical activity are missing.

With regard to sarcopenia (low mass plus low strength or low function), we are not aware of a study of RA patients providing prevalence data. Our results can be appreciated (sarcopenia in 1/6 of RA patients and in 0/52 of HC) however, numbers were too small to allow conclusions. In addition, appropriateness of using handgrip strength as a representative test within the definition of sarcopenia in RA patients is questionable. Inflammation prone joints are typically metacarpophalangeal and proximal interphalangeal, thus disease activity might significantly interfere with test procedure. Our results confirm such a local, mechanical limitation as we saw considerable lower grip strength in RA compared to HC at baseline with no decline over time, while rates of low muscle mass were comparable between groups and declined in RA patients. Furthermore, disease-specific cut-offs are missing. Gait speed is an alternative test in RA patients as a rather nonspecific but accepted test of muscle function in sarcopenia.^15^

With regard to change in muscle mass over time, we found a significant loss of ALMI in RA patients of 0.86 % per year. Studies prospectively examining DXA-derived changes in female ALMI over time are very scarce. A Japanese study of 148 women with RA did not find a significant annual decline of muscle mass. Patients with moderate to high disease activity kept their muscle mass on the same level, whereas those with low disease activity even showed slight gains in ALMI.^13^ However, age range was wide and distribution of participants with regard to menstrual transition period was not defined.

How do our results of muscle mass decline in RA patients compare to a healthy aging trajectory? The Study of women’s health across the nation (SWAN) provided DXA-derived values of total body lean mass of 1’246 peri-menopausal women without RA. Age at final menstrual period was 52.2 (SD 2.8) years. Muscle mass declined in the years from around 8 years prior, till 2 years after the final menstrual period by 0.2 % per year. During an around 8 years follow-up thereafter, a time point comparable to our postmenopausal sample of HCs, no further decline in muscle mass took place.^16^ The comparability to the SWAN cohort is limited by different origins of the participants (SWAN included white, black and Asian women) and muscle mass parameter (SWAN measured whole body lean mass). However, rate of muscle mass decline of our postmenopausal RA patients considerably exceeded the decline of healthy women in SWAN during their menstrual transition period.

In the Health Aging and Body Composition (Health ABC) study, 567 with women showed a comparable muscle mass decline of 0.85 % per year. However, participants were considerably older (73.4 [SD 2.8] years vs. 64 [SD 9.3] years in our study) suggesting an accelerated muscular aging pattern in RA patients. ^17^

With regard to potential causes of accelerated decline of muscle mass in RA patients, we found an inhibition of TNFα to be independently associated with a significant gain in ALMI over time. Vial et al. did not find a differential effect on annual change in muscle mass between TNFα inhibitors versus non-TNFα inhibitory therapy in 83 French RA patients (75% women).^18^ Whereas anti-TNFα users also seemed to benefit with an annual gain in ALMI from 7.7 (SD 1.4) to 7.9 (SD 1.5) Kg/m^2^, comparison to HC did not reveal significant treatment effects over time. In concordance with Vial et al, a deleterious effect of TNFα inhibition on ALMI seems unlikely. In addition, we did not see an effect of IL-6RA on ALMI. This is in contrast to Tournadre et al, where treatment with tocilizumab over 12 months lead to an increase of ALMI from 6.7 (SD 1.4) to 7.2 (SD 1.5) Kg/m^2^.^19^ In our population, the portion of patients on IL-6RA was only 8 percent, thus, differences in patient characteristics, disease activity, glucocorticoid intake and DMARDs do not allow a comparison of these findings.

With regard to grip strength, the main analysis showed a small gain in RA patients, however, with no significant slope differences compared to HC over time. These results might reflect the potential recovery or stability of muscle strength in circumstances with low level of disease activity, glucocorticoid sparing, good clinical response, decrease of inflammatory cytokines such as TNFα and IL-6 and improved physical activity, as key factors for the management of muscle mass and strength. To what extend an insufficient disease control would be responsible for the disadvantageous differential slope of grip strength in patients on csDMARD versus without such treatment remains unclear, significant differences in DAS28(ESR) were not found.

This study contains some limitations. First, we did not have access to detailed physical activity or musculoskeletal therapy plans of either RA patients or HC. Further, we did not systematically assess level of pain. Next, vitamin D substitution were not equal across the studied groups, however, we included the intake into the model. Furthermore, the subgroup analyses in the present study should be viewed as exploratory, since they were underpowered to capture some additional drug class effects. Finally, the findings in the present dataset are restricted to the population of female RA patients after menopause that do not suffer from concomitant metabolic diseases in addition to RA. Substantial additional efforts would be required to assess the validity of these findings in additional populations.

In conclusion, we found postmenopausal RA patients to be at risk for an accelerated decline of muscle mass over time. Except for the use of TNFα inhibitors, which was associated with sparing of muscle mass, neither baseline muscle mass, disease duration or severity, nor any other patient or disease characteristics independently predicted future decline. For clinical care, we recommend to evaluate changes in muscle mass of RA patients on an individual level. Disproportionally high rates of decline need to be captured to take counteractive strategies. Population-based normal values for annual decline of muscle mass and muscle strengths in both healthy elderly and RA patients remain to be determined.

## Data Availability

All data produced in the present study are available upon request to the authors.

## Acknowledgments

We would like to express our gratitude to the study team as well as to the DEXA team of the Department of Rheumatology and Immunology at Inselspital Bern, University Hospital of Bern, Switzerland, for their valuable contribution to this study.

## Competing interests

All authors declare that there are no competing interests with regard to any process of the study.

## Funding

The study was financially supported by the Alfred and Anneliese Sutter-Stöttner Foundation and by the Novartis Foundation for Medical-Biological Research, P.O. Box CH-4002 Basel.

## Notes

### Competing Interest Statement

The authors have declared no competing interest.

### Funding Statement

The study was financially supported by the Alfred and Anneliese Sutter-Stoettner Foundation and by the Novartis Foundation for Medical-Biological Research, P.O. Box CH-4002 Basel.

### Author Declarations

The ethics committee of the Canton of Bern gave ethical approval for this work (approval #BE165/13).

## References

1. Li, T.H., Y.S. Chang, C.W. Liu, et al., The prevalence and risk factors of sarcopenia in rheumatoid arthritis patients: A systematic review and meta-regression analysis. Semin Arthritis Rheum, 2021. 51(1): p. 236–245.

2. Hirani, V., F. Blyth, V. Naganathan, et al., Sarcopenia Is Associated With Incident Disability, Institutionalization, and Mortality in Community-Dwelling Older Men: The Concord Health and Ageing in Men Project. J Am Med Dir Assoc, 2015. 16(7): p. 607–13.

3. Chin, S.O., S.Y. Rhee, S. Chon, et al., Sarcopenia is independently associated with cardiovascular disease in older Korean adults: the Korea National Health and Nutrition Examination Survey (KNHANES) from 2009. PLoS One, 2013. 8(3): p. e60119.

4. Landi, F., R. Liperoti, A. Russo, et al., Sarcopenia as a risk factor for falls in elderly individuals: results from the ilSIRENTE study. Clin Nutr, 2012. 31(5): p. 652–8.

5. Cruz-Jentoft, A.J., G. Bahat, J. Bauer, et al., Sarcopenia: revised European consensus on definition and diagnosis. Age Ageing, 2019. 48(1): p. 16–31.

6. Roos, F., N. Fankhauser, T.H. Collet, et al., Peripheral Volumetric Muscle Area and Total Body Volume in Postmenopausal Women With Rheumatoid Arthritis. J Clin Densitom, 2021. 24(4): p. 613–621.

7. Aletaha, D., T. Neogi, A.J. Silman, et al., 2010 rheumatoid arthritis classification criteria: an American College of Rheumatology/European League Against Rheumatism collaborative initiative. Ann Rheum Dis, 2010. 69(9): p. 1580–8.

8. Martins, J.C., L.F. Teixeira-Salmela, L.A. Castro e Souza, et al., Reliability and validity of the modified sphygmomanometer test for the assessment of strength of upper limb muscles after stroke. J Rehabil Med, 2015. 47(8): p. 697–705.

9. Ngeuleu, A., F. Allali, L. Medrare, et al., Sarcopenia in rheumatoid arthritis: prevalence, influence of disease activity and associated factors. Rheumatology International, 2017. 37(6): p. 1015–1020.

10. Feklistov A D.N., Toroptsova N, s.a.o.i.w.w.r.a. AB0330 Osteoporosis, and A.o.t.R.D. 2018;77:1340.

11. Matsumoto, Y., Y. Sugioka, M. Tada, et al., Change in skeletal muscle mass is associated with lipid profiles in female rheumatoid arthritis patients -TOMORROW study. Clin Nutr, 2021. 40(6): p. 4500–4506.

12. Alkan Melikoğlu, M., Presarcopenia and its Impact on Disability in Female Patients With Rheumatoid Arthritis. Arch Rheumatol, 2017. 32(1): p. 53–59.

13. Santo, R.C., J.M. Silva, P.S. Lora, et al., Cachexia in patients with rheumatoid arthritis: a cohort study. Clin Rheumatol, 2020. 39(12): p. 3603–3613.

14. Vlietstra, L., S. Stebbings, K. Meredith-Jones, et al., Sarcopenia in osteoarthritis and rheumatoid arthritis: The association with self-reported fatigue, physical function and obesity. PLOS ONE, 2019. 14(6): p. e0217462.

15. Cruz-Jentoft, A.J., J.P. Baeyens, J.M. Bauer, et al., Sarcopenia: European consensus on definition and diagnosis: Report of the European Working Group on Sarcopenia in Older People. Age Ageing, 2010. 39(4): p. 412–23.

16. Greendale, G.A., B. Sternfeld, M. Huang, et al., Changes in body composition and weight during the menopause transition. JCI Insight, 2019. 4(5).

17. Goodpaster, B.H., S.W. Park, T.B. Harris, et al., The loss of skeletal muscle strength, mass, and quality in older adults: the health, aging and body composition study. J Gerontol A Biol Sci Med Sci, 2006. 61(10): p. 1059–64.

18. Vial, G., C. Lambert, B. Pereira, et al., The Effect of TNF and Non-TNF-Targeted Biologics on Body Composition in Rheumatoid Arthritis. Journal of Clinical Medicine, 2021. 10(3): p. 487.

19. Tournadre, A., B. Pereira, F. Dutheil, et al., Changes in body composition and metabolic profile during interleukin 6 inhibition in rheumatoid arthritis. J Cachexia Sarcopenia Muscle, 2017. 8(4): p. 639–646.

